# Enhancing Molecular Classifying Accuracy of Pediatric CNS Tumors: A Dual-Classifier Approach Using DNA Methylation Profiling

**DOI:** 10.1101/2025.04.21.25325928

**Authors:** Esra Moosa, Rania Alanany, Shimaa Sherif, Erdener Ozer, Sukoluhle Dube, Ayesha Jabeen, Ian Pople, Davide Bedognetti, Ata Maaz, Ayman Saleh, William Mifsud, Wouter Hendrickx, Christophe M. Raynaud

## Abstract

**Background:** Accurate classification of pediatric central nervous system (CNS) tumors is critical for optimal treatment yet remains challenging due to the limitations of traditional histopathological methods. DNA methylation profiling has gained attention as a promising tool for the molecular classification of CNS tumors. However, despite its potential clinical value, methylation classifiers remain limited to the research settings. This study aims to assess the use of two DNA methylation-based classifiers for CNS tumor diagnostics, with the eventual goal of integrating them into clinical practice and the impact of technical factors such as fixation methods, DNA quantity, and array choice, while exploring the utility of visualization tools (UMAP/t-SNE) and the integration of molecular data for resolving diagnostic ambiguities.

**Methods:** We analyzed 96 pediatric pathology tissue samples, including 75 CNS tumors, 10 with CNS non-tumoral lesions and 11 with non-CNS tumors, performing 130 methylation analyses. DNA from both formalin-fixed paraffin-embedded (FFPE) and fresh frozen (FF) tissues were used for methylation profiling using the Illumina MethylationEPIC V1 and V2 arrays. The performance of two DNA methylation-based classifiers (Heidelberg and NIH) was evaluated by comparing the classification results with histopathological diagnoses. Technical variables that may affect quality such as DNA quantity, extraction method, and sample fixation were also investigated.

**Results:** Both classifiers demonstrated an 88% concordance with histopathological diagnoses in CNS tumors. Methylation profiling refined the histological diagnoses in 54.66% of cases and contributed to molecular subtyping in 52% of CNS tumor cases. The analysis in a small percentage of cases (5.33%) exhibited conflicting diagnoses, emphasizing the need for cautious interpretation and re-evaluation of the cases of uncertainty. Interestingly, both classifiers also identified CNS non-tumor tissues from tumor cases, although they misclassified some normal tissues and malformations as CNS tumors. Technical factors, including DNA quantity and sample fixation, had minimal impact on classifier performance.

**Conclusion:** This study highlights the potential of DNA methylation profiling as a complementary diagnostic tool in pediatric CNS tumor classification, paving the way for its integration into routine clinical practice. To the best of our knowledge, this is the first publication comparing two DNA methylation classifiers in a pediatric CNS tumor cohort. While the classifiers show promise for improving diagnostic accuracy, especially in complex or undiagnosed cases, they should be used as a complementary tool to histopathologic classification. Further research is needed to validate their integration into clinical practice, including refining technical protocols, addressing limitations, and evaluating long-term clinical outcomes.

## Introduction

Accurate diagnosis and molecular classification of central nervous system (CNS) tumors are critical for optimal patient management and treatment. Historically, CNS tumor classification and grading relied primarily on histological assessment, which, while essential, is inherently prone to inter-observer variability and experience. This results in a significant rate of misclassification among pathologists (1). Recognizing the limitations of morphology-based diagnosis, advanced molecular testing has become indispensable, offering critical insights for diagnosis, prognosis, and targeted therapy (2,3). The World Health Organization (WHO) incorporated molecular analyses alongside histological evaluations in its updated CNS tumor classification systems, first in 2016 and later in 2021, listing numerous genetic alterations critical for diagnosis, including many mutations and copy number variations (4).

Among these molecular advancements, DNA methylation profiling has emerged as a powerful and reliable tool for CNS tumor classification. It has proven especially valuable for diagnosing tumors with challenging or atypical morphology and for identifying biologically distinct subtypes within certain tumor families, such as gliomas, ependymomas and medulloblastomas. For instance, ependymomas, though morphologically similar, are now classified into distinct biological types associated with specific anatomic locations and patient populations. Some ependymoma types are defined by genetic alterations such as ZFTA and YAP1 fusions, while others, such as posterior fossa groups A and B, are most accurately distinguished through DNA methylation profiling (5–9). Similarly, medulloblastoma subgroups, initially characterized by transcriptome analysis, are now reliably classified using DNA methylation profiling (10–12).

Moreover, DNA methylation profiling has shown promise in characterizing new tumor classes, such as those previously grouped under primitive neuroectodermal tumors, which have since been revealed to comprise multiple biologically distinct groups with unique genetic drivers. It has also proven effective in reducing inter-observer variability in morphology-based grading systems, such as for meningiomas, where it can predict recurrence risk more reliably than traditional criteria (13–15). Other examples include its utility in distinguishing pleomorphic xanthoastrocytoma and its anaplastic variant, both of which exhibit highly heterogeneous morphology (16,17).

To capitalize on tumor genomic signatures, a machine learning (ML)-based classifier utilizing DNA methylation profiling was pioneered by the German Cancer Research Center and Heidelberg University (Heidelberg classifier). Trained on a dataset of 2801 neuropathological tumor samples, this classifier has been made freely available as a research tool via a web-based portal (18).. However, its application in a clinically validated environment is limited, as it is not maintained under CAP-accredited laboratory conditions.

To the best of our knowledge, this study represents the first comprehensive use of two DNA methylation classifiers within a pediatric central nervous system (CNS) tumor cohort, aiming to evaluate their potential combined integration into clinical diagnostics. Furthermore, the study compares the employment of DNA methylation profiling using Illumina MethylationEPIC V1 and V2 arrays on formalin-fixed paraffin-embedded (FFPE) and fresh frozen (FF) samples tissue samples. Additionally, it investigates the influence of technical variables—including DNA quantity, and sample fixation—on the quality and reliability of the classification process.

## Material and method

### Sample Collection

This retrospective study included overall 96 pediatric patients under 16 years old who received a final pathology diagnosis at Sidra Medicine, Qatar, between January 2018 and December 2024.

As shown in Table 1, there are 75 cases with CNS tumors, 10 with CNS non-tumoral lesions and 11 with non-CNS tumors. Most representative formalin-fixed paraffin-embedded (FFPE) and fresh frozen (FF) samples were selected. All tumor samples consisted of at least 70% of tumor tissue analyzed with the hematoxylin and eosin (H&E) stained slides.

**Table 1:**
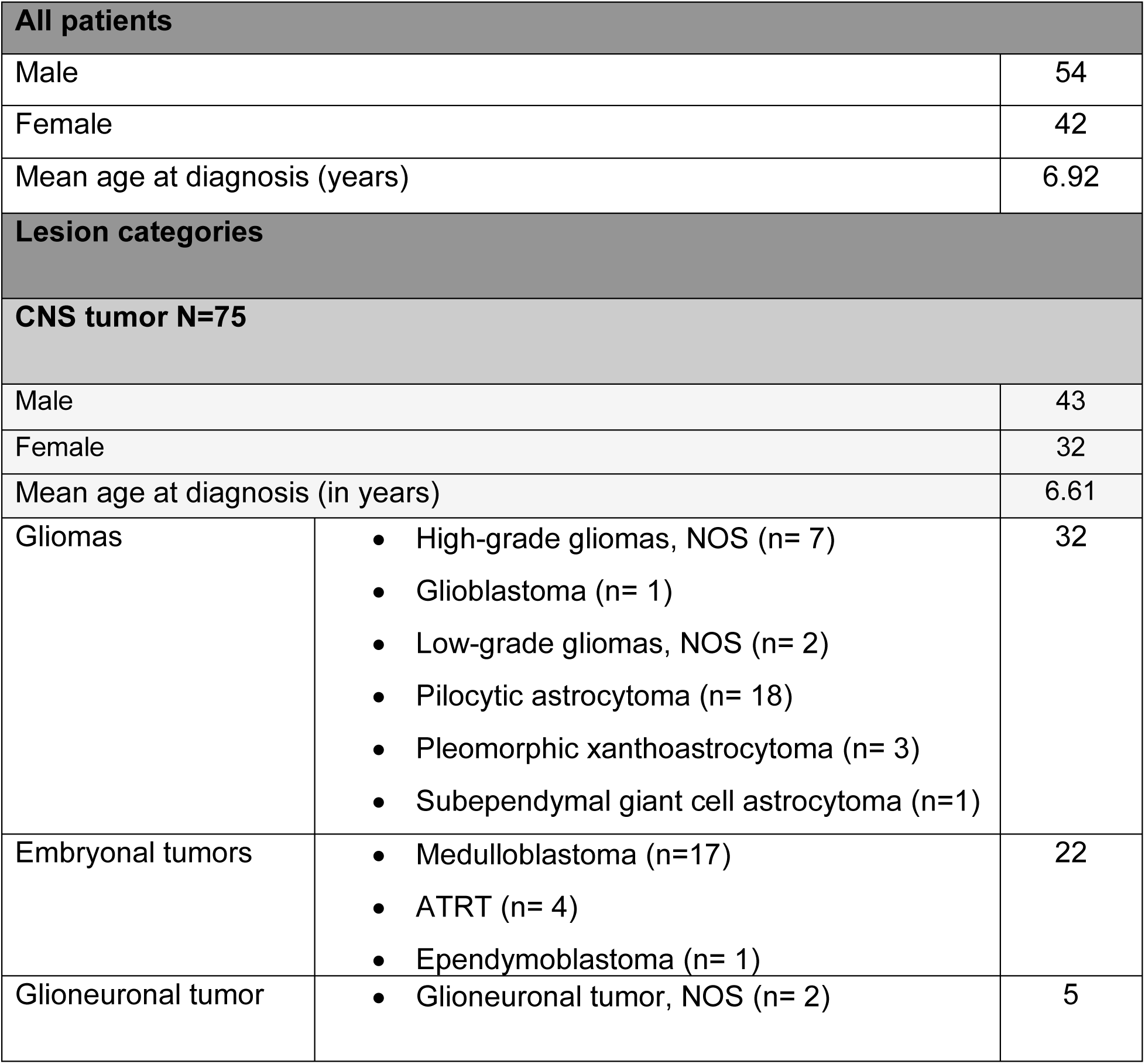
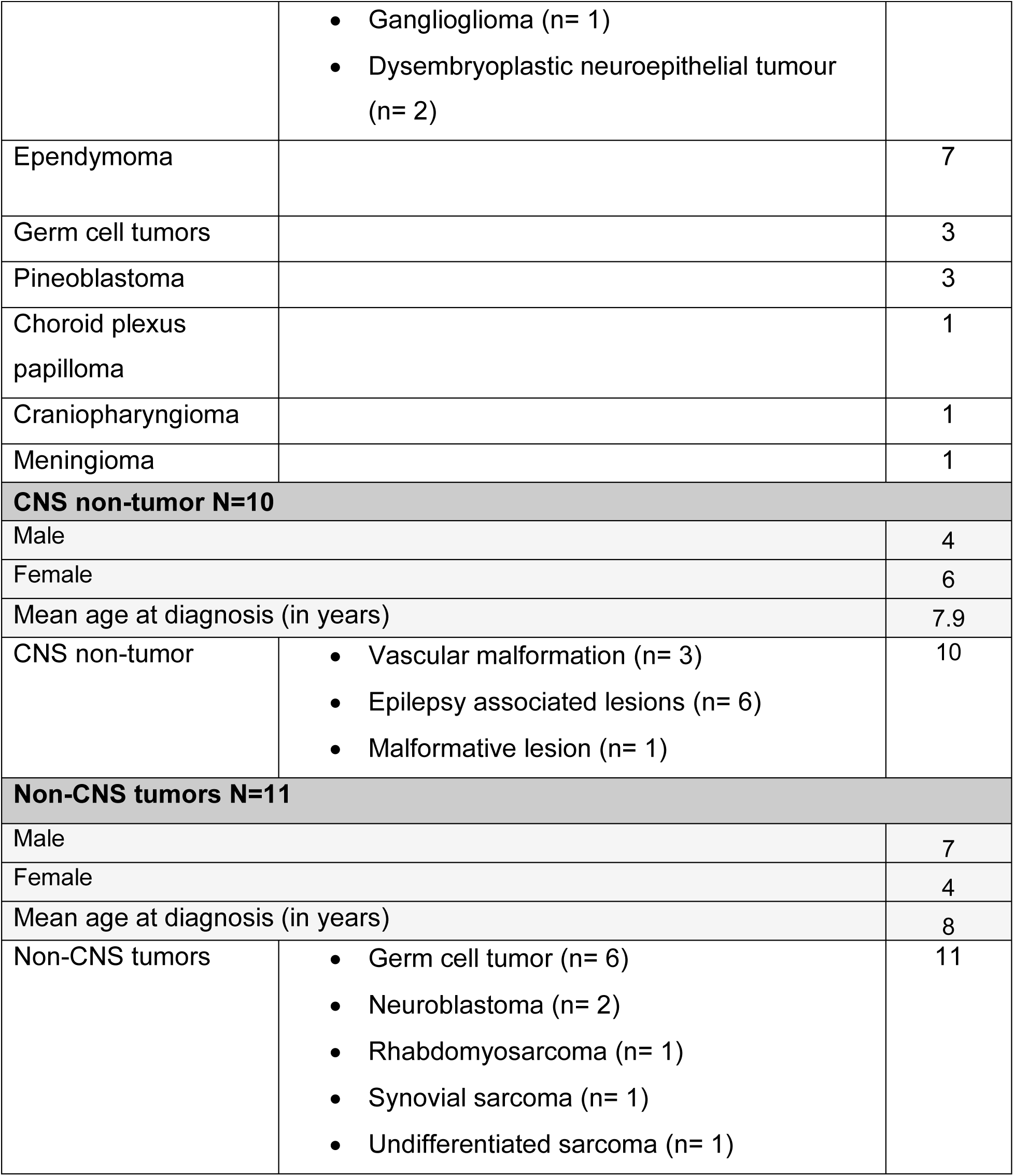
List of patients and lesions characteristics.

### DNA Extraction

Overall, 96 patients’ samples were used for DNA extraction. Nine patients had both FFPE and FF samples, 24 with only FF samples, and 63 with only FFPE samples. A total of 33 DNA extracts were isolated from FF tissues (7 non-CNS tumor and 26 CNS tumors), whereas 72 FFPE samples (4 non-CNS tumors, 10 CNS non-tumors and 58 CNS tumors) were used for DNA extraction.

DNA extraction was performed using either the DNeasy Blood and Tissue Kit (#69504, Qiagen) or the Maxwell® RSC Tissue DNA Kit (#AS1610, Promega) on the Maxwell RSC instrument (#AS4500, Promega) following the manufacturers’ protocols.

Two pathologists (W.M. and E.O.) and one research specialist (C.R.) estimated the number of tumor cells in each sample case. The tumor cell content was assessed based on the H&E stained slides. The H&E slides were cut and pre-checked before submitting the FFPE tissue blocks for DNA extraction and methylation array analysis. DNA was extracted from six 10 µm thickness curls, either immediately or after storage at −80°C for extraction the following day, using the Maxwell® RSC DNA FFPE Kit (#AS1450, Promega).

DNA quantification was performed using the Quantus Fluorometer (#E6150, Promega) with the QuantiFluor® ONE dsDNA System (#E4870, Promega). We aimed to use 500 ng of DNA in 12 µL per sample or the highest possible concentration in up to 35 µL. Bisulfite conversion was carried out using the EZ DNA Methylation™ Kit (#D5002, Zymo Research). Samples with DNA concentrations below 8.5 ng/µL after extraction were excluded (less than 300ng for methylation array analysis).

### Methylation Analysis

In total, we performed 125 methylation array run. Two extracts were analyzed three times as experimental controls and one patient’s DNA samples (from two extractions) was analyzed 17 times as a control in methylation profiling experiments (3 from one extraction and 14 from the second extraction of the same FFPE block).

The DNA sample used as a control in each batch of our analyses served to monitor both the experiment and the technical aspects of the procedure. It was specifically used to ensure that for every batch of 8-46 samples processed at once, the procedure ran smoothly and no technical issues arose, thus validating the accuracy and reliability of the results obtained from each batch.

Methylation data were generated using either the Illumina MethylationEPIC (850k) BeadChip platform (V1) for 40 samples or the Infinium MethylationEPIC v2.0 Kit (930k) (V2) for the remaining samples, following the methodology described by Capper et al. (1) and manufacturer recommendations.

For each sample, paired IDAT files (red and green) were uploaded into the Heidelberg DNA Methylation Tumor Classifier (version 12.8, www.molecularneuropathology.org) or in the methylscape analysis profiler from NIH (www.methylscape.ccr.cancer.gov).

The Heidelberg classifier assigned, when possible, a methylation “superfamily” and, if applicable, a more specific “family”, “class” or “subclass” within that superfamily, along with a calibrated score ranging from 0 to 0.99. This score indicates the degree of similarity between the sample’s methylation profile and the reference database. The scores above 0.9 are considered a match, while scores between 0.9 and 0.3 are considered as no match but indicative as could still be relevant for cases with low tumor content or poor DNA quality. Scores below 0.3 are considered a “no match”. We personally introduced another category with scores between 0.9 and 0.84 as the useful confidence limit was mentioned as possible as low as 0.84 by Capper et al. (18).

With the NIH methylscape classifier, when possible, a methylation “family” and, if applicable, a more specific “Class”are provided along with a calibrated score ranging from 0 to 0.99. Scores above 0.9 are considered a match, while scores between 0.9 and 0.5 are considered as indicative. Scores below 0.5 are considered a “no match”. The NIH methylscape classifier also provides a location of the specific analysed specimen within a UMAP of the samples used for classifier calibration as interpretation can often be improved with visual inspection of unsupervised UMAP embedding.

### t-SNE analysis

Raw signal intensities were extracted from IDAT files using the minfi Bioconductor package (v.11.52.1). Samples were normalized by background correction and dye-bias correction via the funnorm function. Beta values, obtained using the getBeta function, ranged from 0 (completely unmethylated) to 1 (completely methylated). For betas derived from the epic_v2 dataset, the betasCollapseToPfx function from the sesame package (v.1.24.0) was used to collapse the beta values by averaging probes with a common probe ID prefix. Since no batch effects were detected, therefore, no correction was done (data not shown). During the filtration process, the following probes were removed: (a) probes targeting the X and Y chromosomes (n = 11,551), (b) probes containing single-nucleotide polymorphisms within five base pairs of the targeted CpG site (n = 7,998), (c) probes that cross react with more than one region in the human reference genome (hg19) (n = 3,965). Since the publicly available data for training the MNP classifier were analyzed by Illumina Infinium (H450k), while the in-house samples were analyzed using Illumina Infinium Human MethylationEPIC BeadChip (EPIC v1 and EPIC v2). Therefore, to be able to merge all arrays, common probes were used. A total of 358,403 probes were used in downstream analysis. For the unsupervised clustering, the same methods as previously described are used (1). The most variable 32,000 CpGs from all samples (publicly available 2801 samples and 75 in-house samples) were filtered. Eigenvalue decomposition was performed, and 94 nontrivial components were used for t-Distributed Stochastic Neighbor Embedding (t-SNE) analysis. The RSpectra version 0.16.2 and Rtsne version 0.17 were used for the analysis.

### Statistical analysis

Statistical analyses were conducted using GraphPad Prism 10, including calculations for means, standard deviation, t-tests, one way Anova, correlations, and scatter plots.

## Results

### Cohort characteristics

The study included 96 pediatric patients with a mean age of 6.9 years at diagnosis. The cohort comprised 54 males and 42 females. Among the 75 CNS tumor cases, the majority were gliomas (n=32), followed by embryonal tumors (n=22). Non-CNS tumors (n=11) included germ cell tumors, neuroblastomas, and soft tissue sarcomas, while non-tumoral CNS lesions (n=10) included vascular malformations and epilepsy-associated lesions. Detailed demographic and clinical characteristics are summarized in **Table 1**.

### Assay performance of classifiers on the CNS tumors

To evaluate the performance of each classifier we first focused on the CNS tumors and retained only 1 assay per patient. For the Heidelberg classifier, using a calibrated score cutoff of ≥0.9 (as recommended by Capper et al.(1)), the Heidelberg Classifier matched a methylation class at the superfamily level in 79% of patients (59/75) and at a subclass level in 59% of patients (44/75). With a lower cutoff of ≥0.84 (as suggested by Capper et al. (18)), this percentage increased to 84% (63/75) at the superfamily level and 68% (51/75) at the subclass level. (**Figure 1A**). With the NIH methylscape classifier, 92% (69/75) matched a methylation class at the superfamily level and 68% of patients (51/75) matched at the level with a score ≥0.9. (**Figure 1B**).

**Figure 1.**
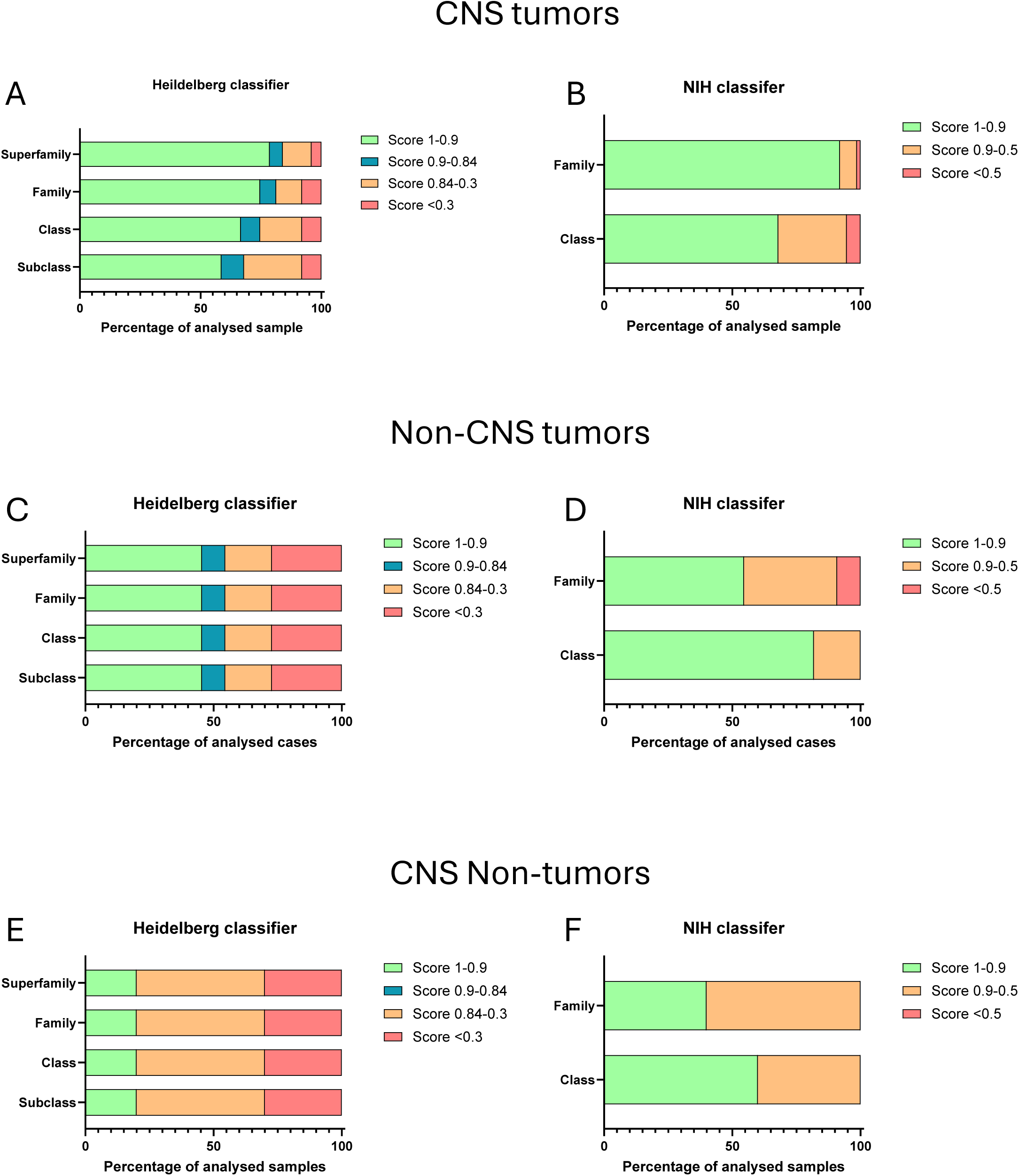
Percentage of the cases with score >0.9 (in green) between 0.9-0.84 (in blue), between 0.84-0.3 (in orange) and <0.3 (in red) at each level of classification with the Heidelberg classifier. For the CNS pediatric tumors (A), for the non-CNS tumors (C) and the CNS non cancer lesions (E). Percentage of the cases with score >0.9 (in green) between 0.9-0.5 (in orange) and <0.5 (in red) at each level of classification with the NIH methylscape classifier. For the CNS pediatric tumors (B), for the non-CNS tumors (D) and the CNS non cancer lesions (F).

While we had a very high concordance between the classification of the 2 classifiers at both superfamily and class level (98.7% and 86.7% respectively) (**Supplementary Figure 1A,B**) Based on the obtained scores by comparing the NIH methylscape classifier to the Heidelberg classifier, the scores obtained at the superfamily and class levels were significantly higher using the NIH classifier (**Supplementary Figure 1C**). It therefore appears that the level of confidence provided by the NIH methylscape classifier is slightly higher than the one provided by the Heidelberg classifier.

### Assay performance of the classifiers on non-CNS tumors and non-tumoral CNS lesions

Additionally, we investigated the possibility of analysis of non-CNS tumors and “CNS non tumors” samples. Yet, to be relevant, the non-CNS tumors analyzed here (germ cell tumors, neuroblastomas, and sarcomas) are all tumor types included in the classifier’s reference database (detailed in **Table1**).

The score obtained with non-CNS tumor were relatively high with 45.5% (5/11) with score >0.9 and 54.5% (6/11) with score >0.84 at the superfamily and subclass level with the Heidelberg classifier (**Figure 1C**). The scores were even higher with 54.5% (6/11) at the superfamily level and even 81.8 (9/11) at the subclass level using the NIH methylscape classifier (**Figure 1D**).

On the other hand the scores obtained on the “CNS non tumors” were comparatively low with 20% (2/10) with score >0.9 at the superfamily and subclass level with the Heidelberg classifier (**Figure 1E**).

Yet, all the ones with score >0.3 confirmed the “normal” condition of the tissue. On the same samples, 40% (4/10) at the superfamily level and even 60% (6/10) at the class level using the NIH methylscape classifier scored >0.9 (**Figure 1F**). Here some of the high scores pointed to a tumor diagnosis while histology revealed no such conditions. The re-evaluation of the H&E confirmed that those were misclassification from the NIH methylscape classifier.

### Evaluation of the impact of technical parameters on assay performance

Both classifiers can be used with DNA isolated from FFPE and FF samples. To investigate whether the use of fresh frozen samples could enhance diagnostic performance by the classifiers we included **9 pairs** of matching fresh frozen and FFPE CNS tumor samples. The number of CpGs detected was trending higher in FF samples (p=0.0562) compared to FFPE samples (**Figure 2A**). But, no statistical difference in the Heidelberg classifier scores (**Figure 2B**) nor in the NIH methylscape classifier scores (**Figure 2C**) was observed at any level of classification (superfamily to subclass). Similarly, when comparing all different CNS tumor FFPE samples analyzed in our cohort (62 samples) with all CNS tumor FF samples (26 samples), we once more observed higher average number of CpGs detected (p=0.0056) in FF samples (**Figure 2D**), but there was once more no statistical difference in both classifiers scores between the two groups at any classification level (**Figure 2E-F**).

**Figure 2.**
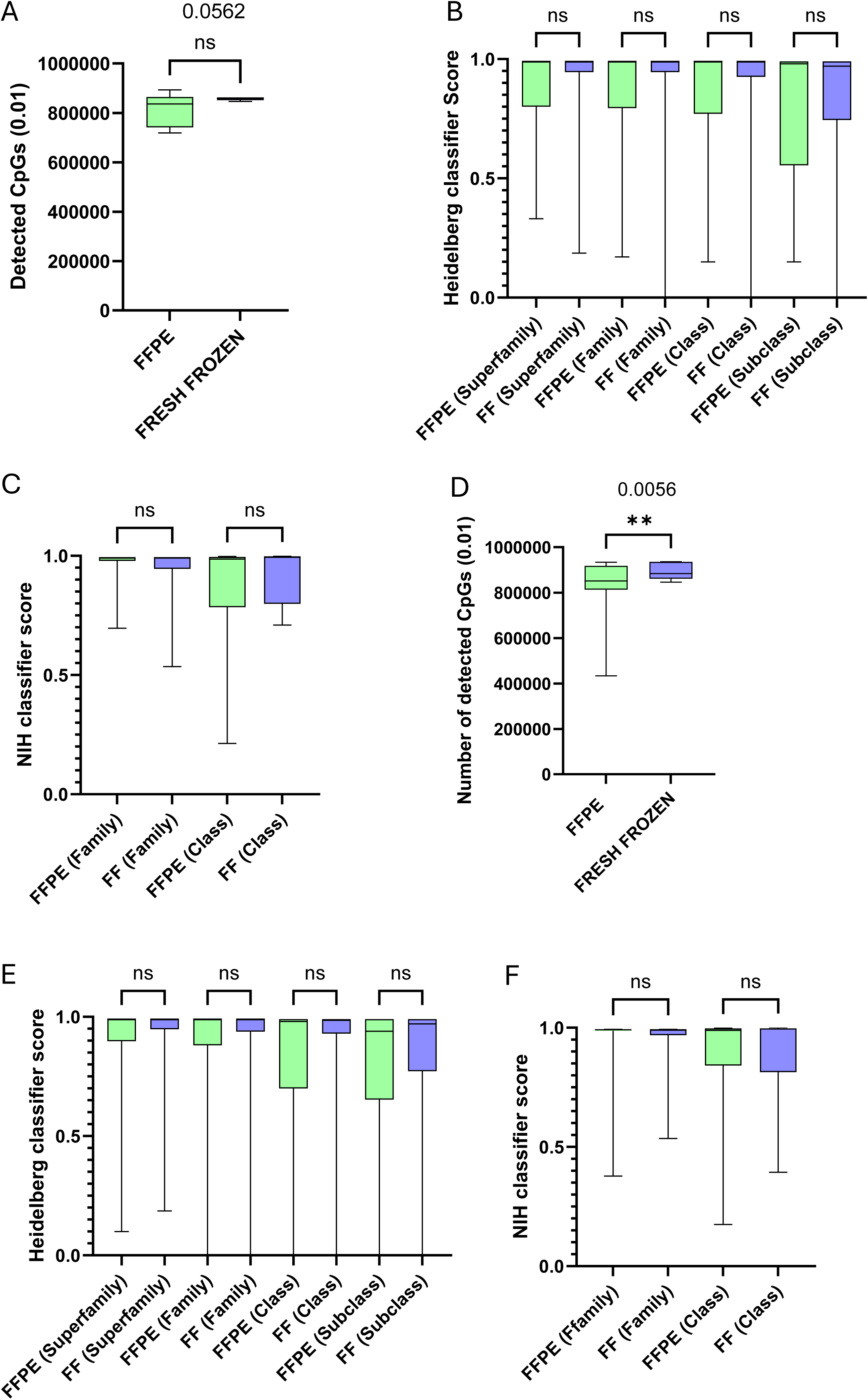
A. Comparison of detected CpGs between paired FFPE and FF sample for the same patients revealed no statistical difference (paired t-test) (p=0.0562). B. Comparison of scores obtained for the 9 pairs of FF and FFPE samples from the same patients at every level of classification of the Heidelberg classifier. The statistical comparison was performed using Anova. C. Comparison of scores obtained for the 9 pairs of FF and FFPE samples from the same patients at every level of classification of the NIH methylscape classifier. (The statistical comparison was performed using Anova. D. Comparison of detected CpGs between all unique FFPE (62 samples) and unique FF (26 samples) CNS tumor samples processed. (unpaired t-test) (p=0.0056). E. Comparison of scores obtained for all the unique FFPE (62 samples) and unique FF (26 samples) CNS tumor samples processed at every level of classification of the Heidelberg classifier. The statistical comparison was performed using Anova. F. Comparison of scores obtained for all the unique FFPE (62 samples) and unique FF (26 samples) CNS tumor samples processed at every level of classification of the NIH methylscape classifier. The statistical comparison was performed using Anova.

Finally, a T-SNE analysis of the samples based on FFPE vs. FF segregation was performed and showed no clustering effect based on whether samples were FFPE or FF, reinforcing the fact that both can be used for analysis and highlighting the robustness of methylation profiling across different sample types (**Supplementary Figure 2**).

We also explored whether the number of analyzable CpGs was correlated with the score obtained. We identified a weak positive correlation between the analyzed CpGs number and the Heidelberg classifier scores at some classification level (p=0.0082, R^2^=0.078 for scores at the superfamily level, p=0.54 R^2^=0.0042 for scores at the family level, p=0.022, R2=0.060 for scores at the class level and p=0.010, R2=0.074 for score at the subclass level, respectively) (**Supplementary Figure 3 A-D)**).

Similarly, with the NIH methylscape classifier, the correlation between the analyzed CpGs number and the Heidelberg classifier scores was not significant at the superfamily level (P=0.90, R2= 0.00018) but was at the class level (p=0.018, r2=0.063) (**Supplementary Figure 3 E-F**). Despite these correlations the plots of CpGs with scores shows that a higher number of analyzable CpGs does not guarantee a higher score with neither of the classifiers, even at the superfamily level of classification (**Figure 3A** and **3B**).

**Figure 3.**
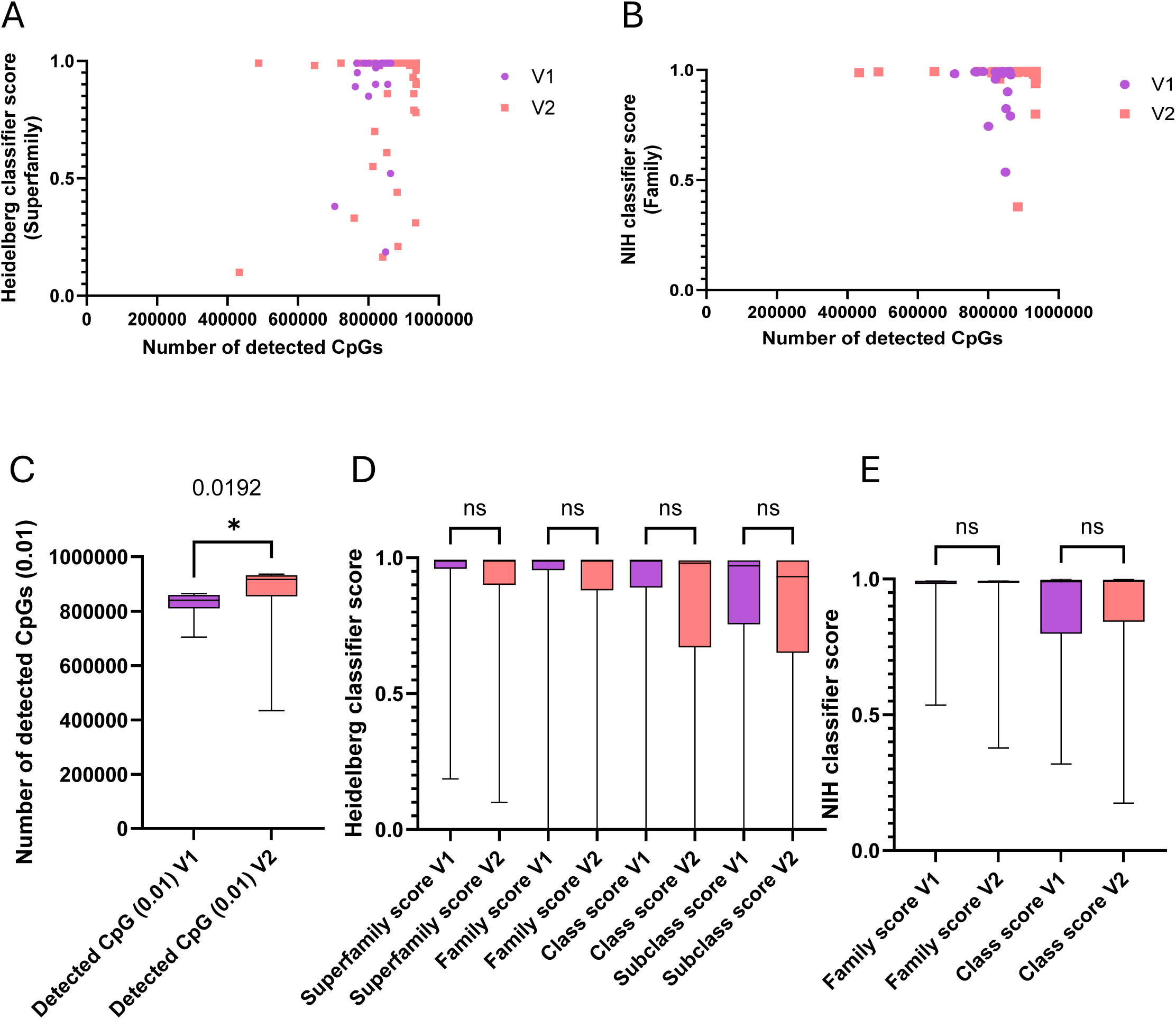
A. Dot plot of the number of detected CpGs with corresponding Heidelberg score at the superfamily level. In violet are the samples processed using Illumina MethylationEPIC v1.0 Kit ( 20 FFPE and 13 FF samples) and in red are the samples processed using Illumina MethylationEPIC v2.0 Kit ( 42 FFPE and 13 FF samples). B. Dot plot of the number of detected CpGs with corresponding NIH methylscape score at the superfamily level. In violet are the samples processed using Illumina MethylationEPIC v1.0 Kit) and in red are the samples processed using Illumina MethylationEPIC v2.0 Kit. C. Comparison of the numbers of detectable CpGs on samples using V1 or V2 Illumina MethylationEPIC kits. Significantly more CpGs were detected with V2 kits (p=0.0192, Unpaired t-test). D. Comparison of scores obtained for all the samples processed with Illumina MethylationEPIC V1 (33 cases) and V2 (55 cases) CNS tumor samples at every level of classification of the Heidelberg classifier. The statistical comparison was performed using Anova. Comparison of scores obtained for all the samples processed with Illumina MethylationEPIC V1 (33 cases) and V2 (55 cases) CNS tumor samples at every level of classification of the NIH methylscape classifier. The statistical comparison was performed using Anova.

Since we used both the **Illumina 850k BeadChip (V1)** in 2023 and the **930k BeadChip (V2)** in 2024, we distinguished between the two bead chip types. We found that, as expected, more analyzable CpGs were recovered with the V2 chip than with V1 (p=0.019) (**Figure 3C**). However, since the CpGs used for classification were defined among the 850k CpGs present in V1 (and still present in V2), the additional CpGs recovered with V2 did not improve the average scoring with either classifier at any level of classification (**Figure 3D-E**).

Finally, despite efforts to optimize DNA extraction and using the recommended 500ng by Illumina for EPIC analysis, we were unable to consistently recover the required amount of DNA. In some cases, we performed analysis with as little as 300ng, which became our internal cutoff (above the 250ng recommended by illumina). Interestingly, using the recommended DNA amount did not guarantee a high number of CpGs recovered or a good score. Conversely, using low DNA quantities (as low as 300ng) could still result in a good number of CpGs detected and a DKFZ score ≥0.9 (**Supplementary Figure 4A-E**).

### Assay reproducibility

We included one particular sample (a medulloblastoma) at each Illumina MethylationEPIC experiment as an internal control across the 17 separate experiments we ran. While the score obtained at the superfamily and family level were nearly identical across all 17 experiments some score discrepancies were seen at the class and subclass level with the Heidelberg classifier (**Supplementary Figure 5A**). This minor variation observed in the Heidelberg score was not seen with the NIH methylscape classifier (**Supplementary Figure 5B**). These minor variations were not linked with the freeze thaw cycles of the sample nor was the number of CpGs detectable (**Supplementary Figure 5C**). Similarly, two additional samples were run respectively 3 times and had very similar results at each run with both classifiers (**Supplementary Figure 5D-I**).

### Primary tumor and relapse comparison

One patient with atypical teratoid/rhabdoid tumor underwent 3 surgeries for a primary tumor and 2 relapses. All 3 analysis (FFPE Samples) gave a similar diagnostic from the classifiers with scores ≥ 0.84 and >0.9 respectively (**Supplementary Figure 6 A-B**). Interestingly the diagnostic of the primary tumor had a score slightly lower than the relapse, but that might be only due to the purity of the cancer tissue presents within the selected block. Similarly, 2 other patients with Ependymomas (both frozen samples) and adamantinomatous craniopharyngioma (both FFPE samples) had primary and relapse analyzed. Like for the previous example both primary and relapse returned the same classification with similar score with the Heidelberg classifier (**Supplementary Figure 6 C-F**). For the Ependymomas a lowers score (<0.5) at the class level with the NIH classifier requalified the subclass 1E as subclass 1D leading to a minor discrepancy between the classifiers.

### Diagnostic performance of the classifiers

We observed a very high concordance in the classification between the two classifiers. Indeed, within the CNS tumors, the 2 classifiers match together at the superfamily level in 98.66% of cases (74/75) of cases and in 86.66% (65/75) of cases at the class level (**Supplementary Figure 1 A-B**). The discrepancy observed at the Class level were often very minor such as Posterior Fossa Group A (pfa) Ependymoma, Subclass 1b for the Heidelberg classifier versus Posterior Fossa Group A (pfa) Ependymoma, Subclass 1c for the NIH classifier. The rest of the discordance at the class level were due to very low score obtained in either or both classifiers. The scores obtained with the NIH classifier were slightly higher at the superfamily level (p=0.0008), and class level (p=0.008) between the two classifiers (**Supplementary Figure 6 C**).

We then assess the correspondence between histology diagnostic and the classifiers prediction for each of the 75 patients. We observed that in 88% (66/75) of cases the classifier confirmed the diagnostic posed by the initial pathological diagnosis (based on histology alone), and in 54.7% (41/75) the classifier provided a more refined diagnosis than the one identified with histology alone (**Figure 4A**). In only 12% (9/75) we obtained a conflicting diagnosis between histology and the classifiers.

**Figure 4.**
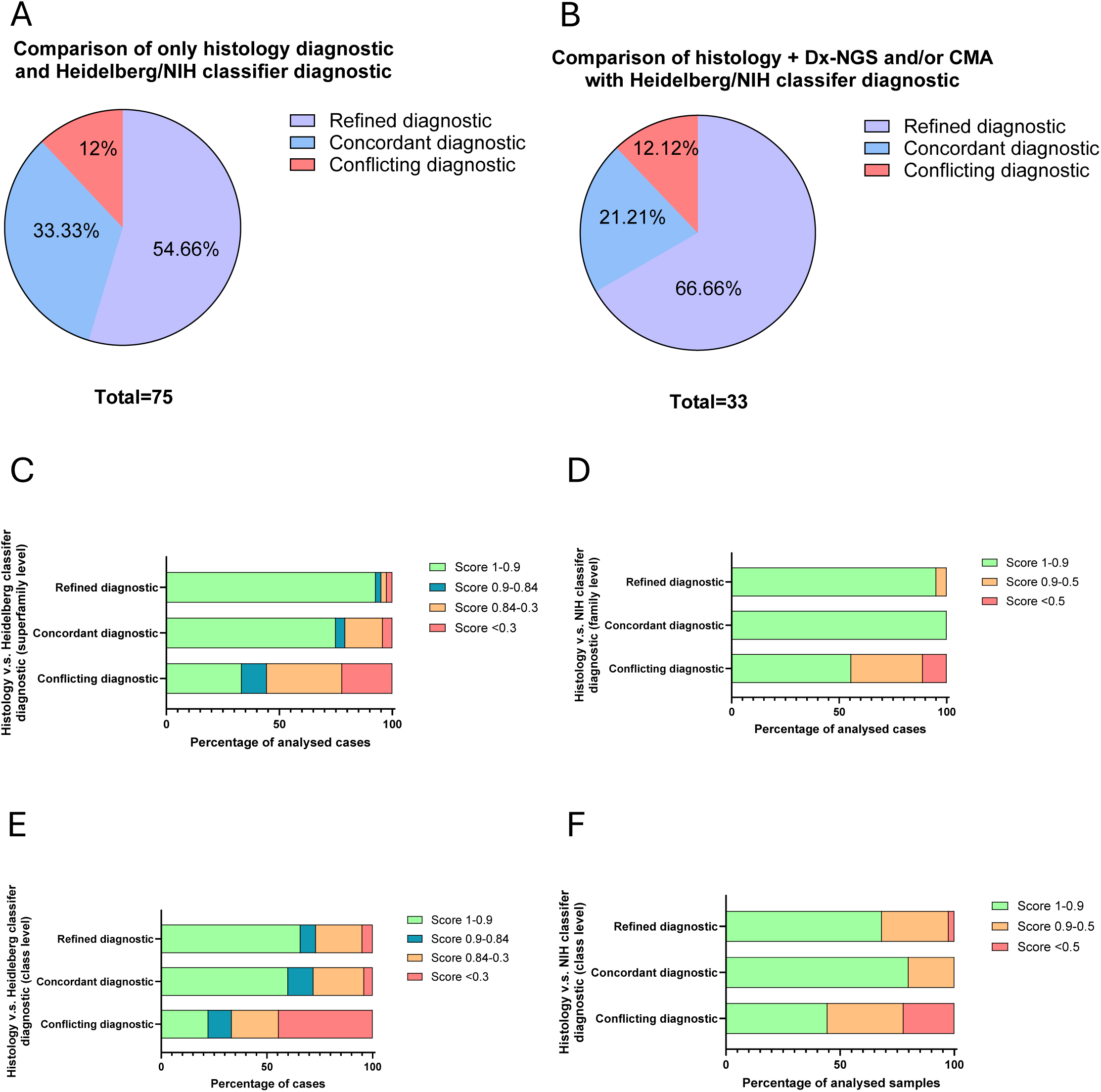
A. Pie chart of the percentage of the 75 CNS tumors with refined (purple), concordant (blue) and conflicting (red) diagnostic provided by the classifiers compared to the histology diagnostic alone. The numbers are identical with both classifiers; therefore the same pie chart is representative for both classifiers. B. Pie chart of the percentage of the 33 CNS tumors with refined (purple), concordant (blue) and conflicting (red) diagnostic provided by the classifiers compared to the histology diagnostic complemented with Dx-NGS and CMA data. C. Percentage of the 75 CNS tumors cases with score >0.9 (in green) between 0.9-0.84 (in blue), between 0.84-0.3 (in orange) and <0.3 (in red) for each concordance between histology only diagnostic and Heidelberg classification (Refined, concordant and conflicting) at the superfamily level (C) and the subclass level (E). D. Percentage of the 75 CNS tumors cases with score >0.9 (in green) between 0.9-0.5 (in orange) and <0.5 (in red) for each concordance between Histology only diagnostic and NIH methylscape classification (Refined, concordant and conflicting) at the superfamily level (D) and the subclass level(F).

Among the 75 cases we obtained diagnostic next generation sequencing (Dx-NGS) and/or chromosomal microarray analysis (CMA) for 33 patients. Among these patients both classifiers provided matching diagnostic in 84.9% (28/33) of cases with the diagnostic obtained with histology and molecular analysis and the classifiers diagnostic. In 15.1% (5/33) we had discordant diagnostic (**Figure 4B**). Surprisingly, even with the integration of next-generation sequencing (NGS), the proportion of cases in which the classifier provided a more refined diagnosis was higher than the global average (66.6% vs. 54.65%, respectively). This discrepancy may reflect the inclusion of more complex cases that necessitate advanced molecular characterization.

Besides, the score obtained with Heidelberg and NIH classifier in the concordant and refined diagnostic, at the superfamily and subclass level, were usually high compared to the conflicting ones (**Figure 4C-D**). The conflicting diagnostics were most often due to low scores.

Among the 75 CNS tumor cases, 9 (12%) exhibited conflicting diagnoses between histopathology and the methylation classifiers (**Figure 4A**). Four cases had low classifier scores (≤0.84 for Heidelberg and ≤0.5 for NIH), with two classified as normal tissue, likely due to low tumor content in the analyzed FFPE blocks despite representing >70% of the tissue on H&E slides. The remaining five cases had high classifier scores (≥0.84 for Heidelberg and ≥0.9 for NIH) but conflicted with histopathological diagnoses, which were confirmed upon H&E reinspection and additional molecular testing (Dx-NGS) (**Figure 4B**) and are therefore true conflicts.

### UMAP/t-SNE Analysis

We utilized the online database and script published by Capper et al. to generate t-SNE plots and also employed the UMAP provided by the NIH classifier for clustering analysis of our methylation data. **Figure 5A** presents the t-SNE plot for our 75 CNS tumor samples mapped against 2801 samples used to create the Capper et al. classifier in their first publication (1). Most samples were clustered within the expected subtype group; however, several outliers were observed, indicating discrepancies between histology and classification results. Most but not all outliers were of low scores, and the analysis of the UMAP/t-SNE could shed light onto the classification of those samples and their scores. For example, one such outlier was a Pleomorphic xanthoastrocytoma (PXA) sample, classified as low-grade Ganglioglioma by both classifiers, despite histological diagnosis of PXA. In the NIH UMAP (**Figure 5B**), this sample clustered with the Ganglioglioma group, explaining its high NIH classifier score. However, in the t-SNE plot (**Figure 5C**), it clustered between Infantile High-Grade Glioma (IHG) and low-grade glioma ganglioglioma (LGG-GG), showing a Heidelberg classifier score of 0.86 at the superfamily level and 0.77 at the subclass level.

**Figure 5.**
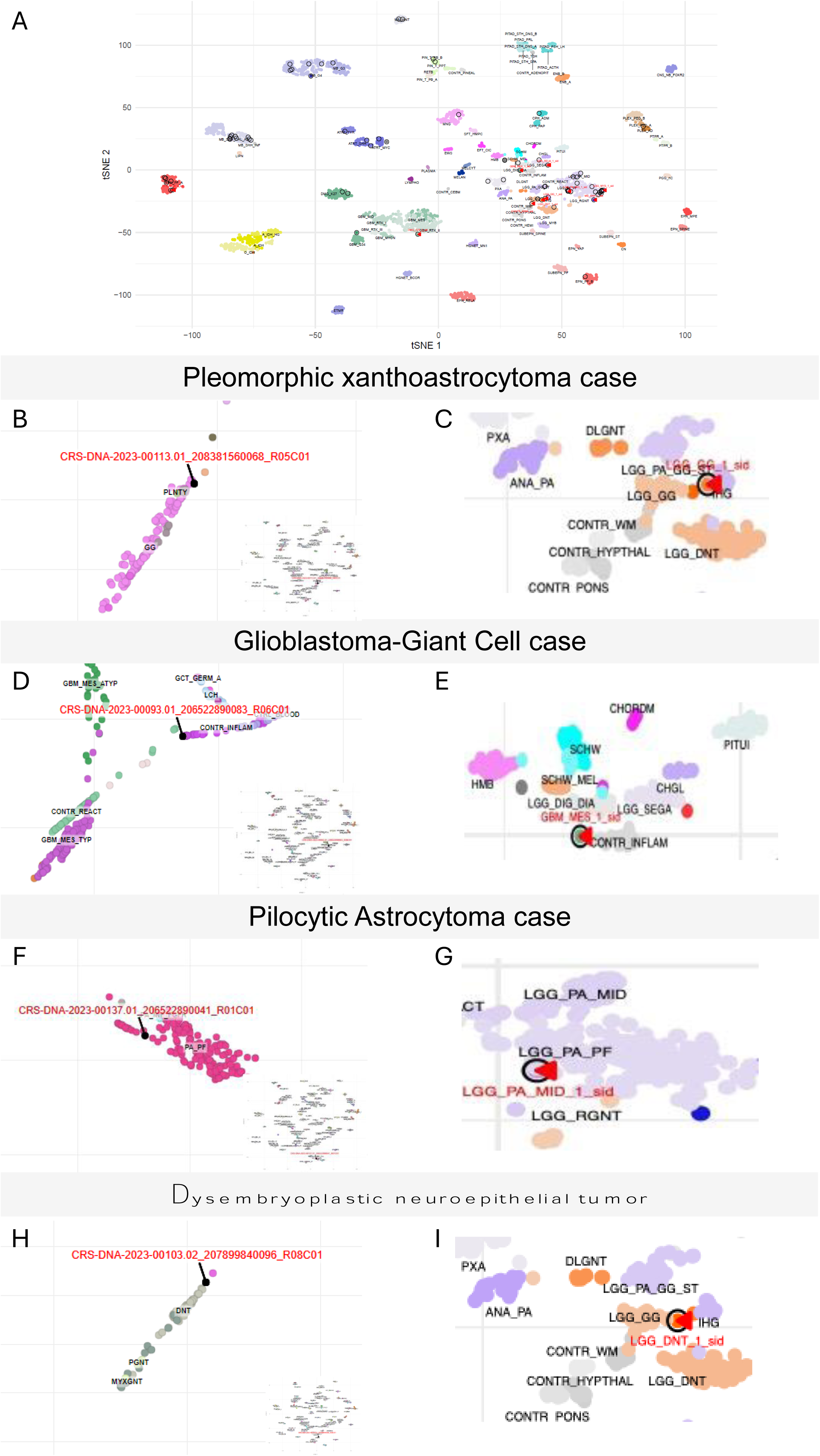
A. t-SNE representation of all the 75 CNS tumors (circled in red) against the 2801 samples published in Capper et al. Our samples are circled in black and the samples not falling in their designated category (outliers) are indicated with a red arrow. Focused localization on the Pleomorphic xanthoastrocytoma case on the NIH classifier of UMAP (B) and focus around this case on the t-SNE performed in our laboratory (labeled in red as LGG_GG_1_sid on the graph) (C). Polymorphous low-grade neuroepithelial tumor (PLNTY) and Ganglioglioma (GG) in the NIH classifier (B). Low-grade glioma, ganglioglioma, (LGG_GG), Infant High-Grade Gliomas (IHG) in the t-SNE (C). Focused localization of the Glioblastoma-Giant Cell case on the NIH classifier of UMAP (D) and focus around this case on the t-SNE performed in our laboratory (labeled in red as GBM_MES_1_sid on the graph) (E). Control inflammatory (CONTR_INFLAM), Langerhans cell histiocytosis (LCH), Glioblastoma, IDH-wt, mesenchyma subtype, atypical (GBM_MES_ATYP) in the NIH classifier (F). Subependymal giant cell astrocytoma (LGG_SEGA), control inflammatory (CONTR_INFLAM), desmoplastic infantile ganglioglioma (LGG_DIG), Schwannoma (SCHW_MEL) in the t-SNE (E). Focused localization on the Pilocytic Astrocytoma case on the NIH classifier of UMAP (F) and focus around this case on the t-SNE performed in our laboratory (labeled in red as LGG_PA_MID_1_sid on the graph) (G). Pilocytic astrocytoma, posterior fossa (PA_PF), infratentorial pilocytic astrocytoma subclass FGFR1-altered (PA_INF_FGFR) in the NIH classifier. Pilocytic astrocytoma, posterior fossa subclass (LGG_PA_PF), MC rosette-forming glioneuronal tumor (LGG_RGNT) in the t-SNE (G). Focused localization on the Dysembryoplastic neuroepithelial tumor case on the NIH classifier of UMAP (H) and focus around this case on the t-SNE performed in our laboratory (labeled in red as LGG_DNT_1_sid in the graph) (I). Dysembryoplastic neuroepithelial tumor (DNT), Papillary glioneuronal tumors (PGNT) in the NIH classifier (H). Low-grade glioma, ganglioglioma, (LGG_GG), Infant High-Grade Gliomas (IHG), control tissue (CONTR_WM), Dysembryoplastic neuroepithelial tumor (LGG_DNT) in the t-SNE (I).

Another outlier, a Glioblastoma-Giant Cell, showed high scores at the superfamily and class levels but lower subclass scores. The NIH UMAP clustered this sample with Control Inflammation (**Figure 5D**), suggesting potential issues with sample purity or contamination. The t-SNE plot for the same sample (**Figure 5E**) further separated this sample from any distinct cluster.

A Pilocytic Astrocytoma (PA) demonstrated a discrepancy between the Heidelberg and NIH classifiers (at the subclass level), with the Heidelberg classifying it as PA, Midline. The NIH classifier providing a high-confidence score than the Heidelberg one. Both NIH UMAP and our t-SNE clustered this sample within the PA_PF (Pilocytic Astrocytoma, Posterior Fosa) group (**Figure 5F-G**).

A rare DNT sample was correctly identified by both classifiers despite low scores. The NIH UMAP but not t-SNE clustering confirmed the diagnosis (**Figure 5H, I**), increasing confidence in the result for the NIH classifier.

## Discussion

This study demonstrates that DNA methylation profiling (MP) has significant potential for CNS tumor diagnostics, particularly in pediatric cases. Both classifiers evaluated showed an 88% concordance with histopathological diagnoses, aligning with findings from Capper et al. and results from other centers (49–95%) (1,8,9,21). Methylation profiling provided diagnostic refinement in 54.7% of cases, highlighting its ability to uncover new pediatric brain tumor subtypes. However, ever since the WHO’s 2021 recommendation of methylation classifiers for CNS tumor subtyping (22,23), much progress is need to bring these research tools into clinical practice, underscoring the need to bridge the gap between their clinical potential and current status.

Our analysis confirmed that technical factors such as fixation method (FF vs. FFPE), DNA quantity, and the technical update from Illumina MethylationEPIC V1 to V2 arrays had minimal impact on classifier performance, consistent with previous studies (8,24). FF samples, which generally offer better DNA quality, performed similarly to FFPE samples under our specific conditions, suggesting no significant improvement in classification when using FF samples. DNA quantity alone was not a reliable predictor of assay performance, as even low-yield samples could achieve sufficient CpG detection and classifier accuracy, while high DNA concentration did not necessarily ensure optimal results. Furthermore, the change between EPIC V1 or V2 arrays did not significantly impact classification scores either, indicating that both versions are suitable for clinical implementation.

Despite these advances, challenges remain. Lower scores were observed for non-tumor CNS lesions, and the NIH Methylscape classifier occasionally produced false positive tumor classifications, emphasizing the need for cautious interpretation in non-cancerous cases. These discrepancies highlight the importance of ensuring adequate cancer cell content for methylation profiling of tumor tissue and underscore the complementary role of classifiers alongside histopathology. The integration of molecular data (e.g., Dx-NGS) and visualization tools (e.g., UMAP/t-SNE) proved invaluable for resolving diagnostic ambiguities, particularly in cases with low tumor content or high biological complexity. Our study documents several cases that highlight the critical role of integrating UMAP/t-SNE analysis with methylation profiling and histopathology to enhance diagnostic accuracy, especially in complex or rare tumor types. For instance, the discrepancies observed in the Pleomorphic Xanthoastrocytoma (PXA) case underscore the challenges in diagnosing morphologically heterogeneous tumors and emphasize the necessity of combining histopathological review with methylation profiling. The Glioblastoma-Giant Cell case demonstrates how UMAP/t-SNE can identify technical limitations that may influence classifier performance, providing valuable insights for optimizing diagnostic workflows. Similarly, the Pilocytic Astrocytoma (PA) case illustrates the importance of using multiple classifiers and visualization tools to resolve ambiguities in tumor subclassification. Finally, the Dysembryoplastic Neuroepithelial Tumor (DNT) case showcases the utility of methylation profiling and UMAP/t-SNE in diagnosing rare tumors, even when classifier confidence scores are low.

Reproducibility is critical for clinical integration. In our study, the use of inter-experiment controls—specifically a designated control sample—helped identify a technical issue in one batch of analyses, where the control sample showed significantly lower CpG detection and failed to reach a score ≥0.9. This underscores the importance of internal controls and the inclusion of technical parameters, such as DNA quantity, bisulfite conversion quality, and detected CpG sites, in diagnostic reports to facilitate accurate interpretation of results. Additionally, the batching requirement for processing samples in groups of eight caused delays in diagnostic turnaround time, prompting exploration of alternative methods like Oxford Nanopore Technology sequencing, which could improve workflow efficiency.

While the classifiers showed high concordance with histopathology when scores were ≥0.9, they should not replace traditional methods but rather serve as complementary tools to confirm diagnoses or flag cases for re-evaluation. In 5.3% of cases (4/75) with scores >0.9, reclassification occurred, a rate slightly lower than the 6–25% reclassification rates reported by Capper et al. (18). In these cases, the original histological diagnoses remained intact upon further review, including H&E re-evaluation and Dx-NGS results. Unsupervised plotting did not resolve these discrepancies, reinforcing the need for a multidisciplinary approach to CNS tumor diagnostics. Furthermore, the classifiers showed potential utility in certain non-CNS tumor subsets, though they struggled to distinguish normal tissue from cancerous, highlighting the need for cautious application.

Several limitations warrant further investigation, including the effect of sample age on methylation profiling and the need for clinical follow-up data. Additionally, the integration of MP results into multidisciplinary team discussions, as recommended by the Haarlem and ICCR guidelines, is essential for comprehensive diagnostic evaluations. While MP holds significant promise for CNS tumor diagnostics, its routine clinical use requires continued refinement of technical processes, validation of reproducibility, and further research into its clinical applicability. Ultimately, methylation profiling should serve as an adjunct to traditional methods, guiding clinicians in challenging cases and aiding in the identification of new tumor subtypes.

## Abbreviations

DNA: Deoxyribonucleic acid
FFPE: Formalin-Fixed Paraffin-Embedded
FF: Fresh frozen
MP: Methylation profiling

## Declarations

### Ethics approval and consent to participate

All samples were used under IRB #150900 approved by Sidra IRB committee in accordance with Qatari Regulations, Sidra Medicine policies and procedures and all applicable regulations related to human research protection. Some patients were not consented and sample were anonymized, therefore human Ethics and Consent to Participate declarations is not applicable.

## Data Availability

All data produced in the present study are available upon reasonable request to the authors

## Acknowledgement

We would like to thank the genomic core of Sidra medicine for it’s help in this manuscript and the Illumina MethylationEPIC analysis.

## Consent for publication

All authors consent for publication of results

## Availability of data and materials

The datasets generated and/or analyzed during the current study are available in the supplementary table 1.

## Author’s contribution

W.M. and E.O. selected all the samples, E.M. and S.D. cut all the samples, E.M., C.R., performed the experiments, C.R., W.M., E.O., R.A. and E.M. performed the analysis C.R., drafted the manuscript and the figures, W.H., S.S., A.M., A.S., D.B., I.P. and E.O. edited the text. Conceptualization by W.M., C.R., E.O. and W.H., C.R. project supervision and coordination. All authors reviewed the manuscript.

---

**Supplementary Figure 1.**
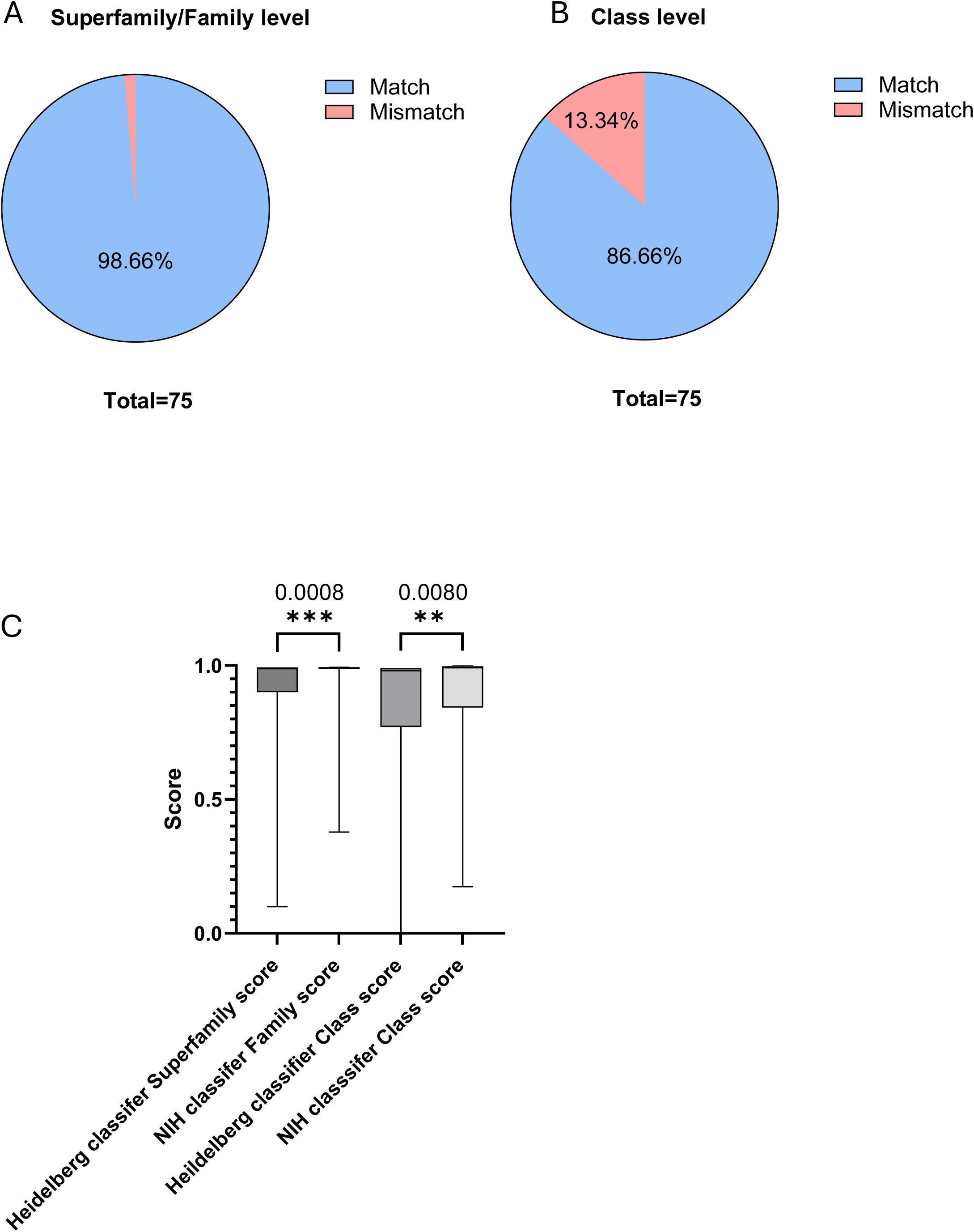
Pie chart of the percentage of the 75 CNS tumors concordance of classification between Heidelberg and NIH methylscape classifiers at the superfamily (Heidelberg)/family (NIH) level (A) and at the Class level (B). C. Comparison of the scores obtained at the superfamily level and class level with both Heidelberg and NIH classifiers. Significantly higher scores were obtained with NIH classifier at the superfamily (Heidelberg)/family (NIH) (P=0.0008, paired t-test), and class level (P=0.008).

**Supplementary Figure 2.**
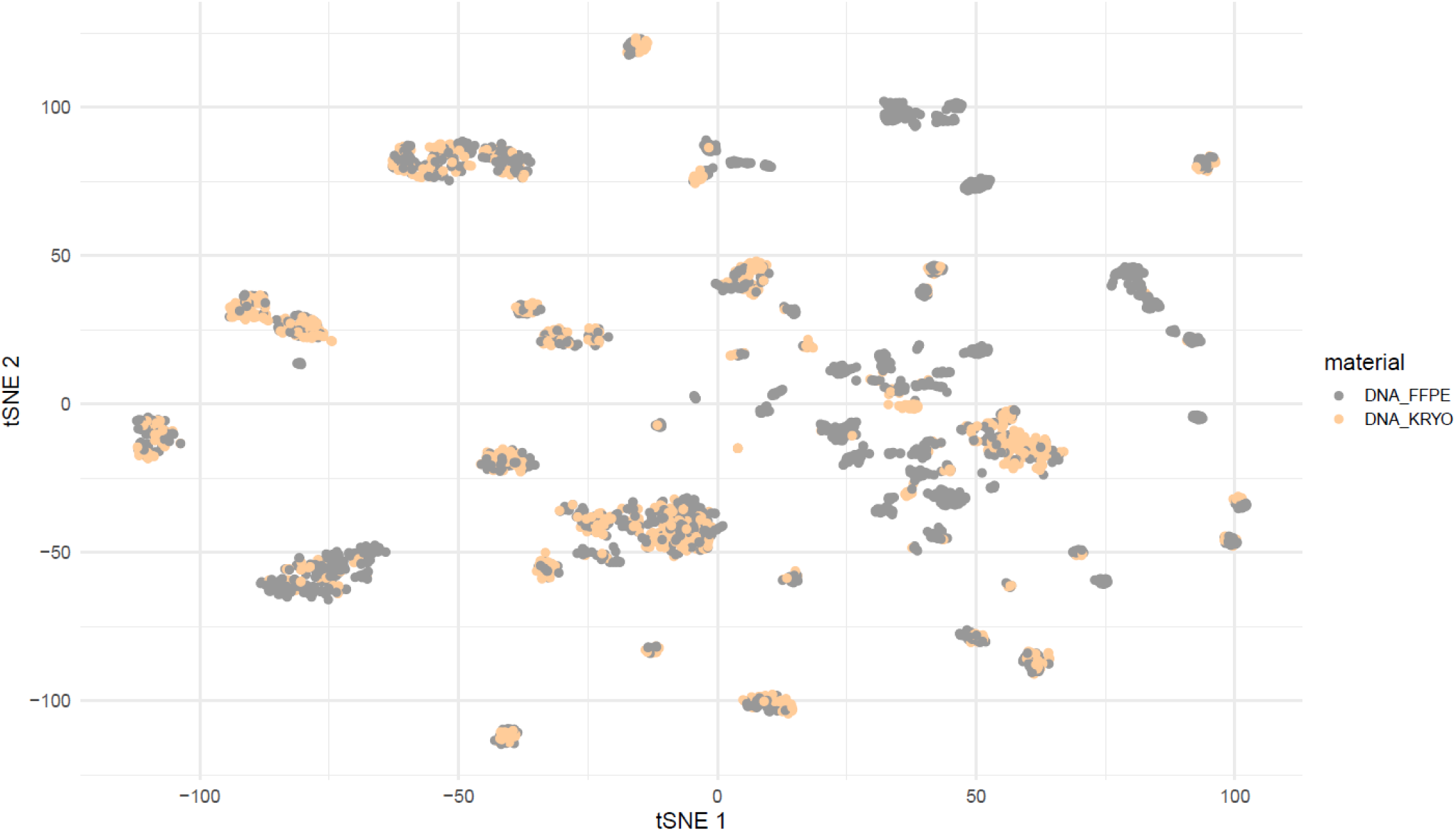
UMAP (Uniform Manifold Approximation and Projection) visualization of DNA methylation profiles from FFPE and FF samples. The plot demonstrates no significant clustering based on sample type (FFPE vs. FF).

**Supplementary Figure 3.**
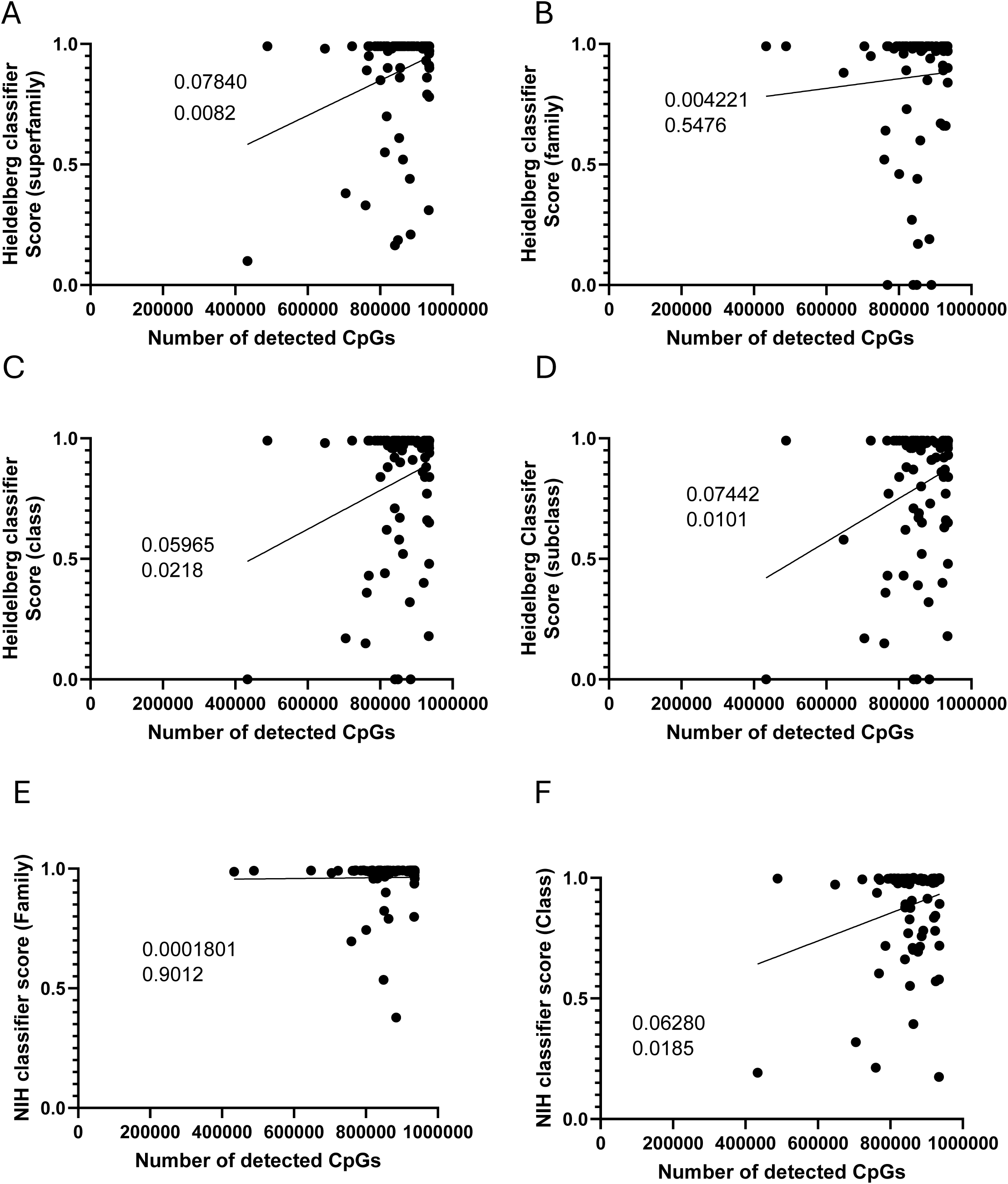
Dotplot of 88 samples (duplicates excluded) detectable CpGs against the Heidelberg score at the superfamily (A), Family (B), Class (C) and Subclass level (D). Linear regression with R^2^ (up) and P value (down) are provided for each. Dotplot of 88 samples (duplicates excluded) detectable CpGs against the NIH methylscape score at the family (E), and Class level (F). linear regression and P value and R^2^ are given for each.

**Supplementary Figure 4.**
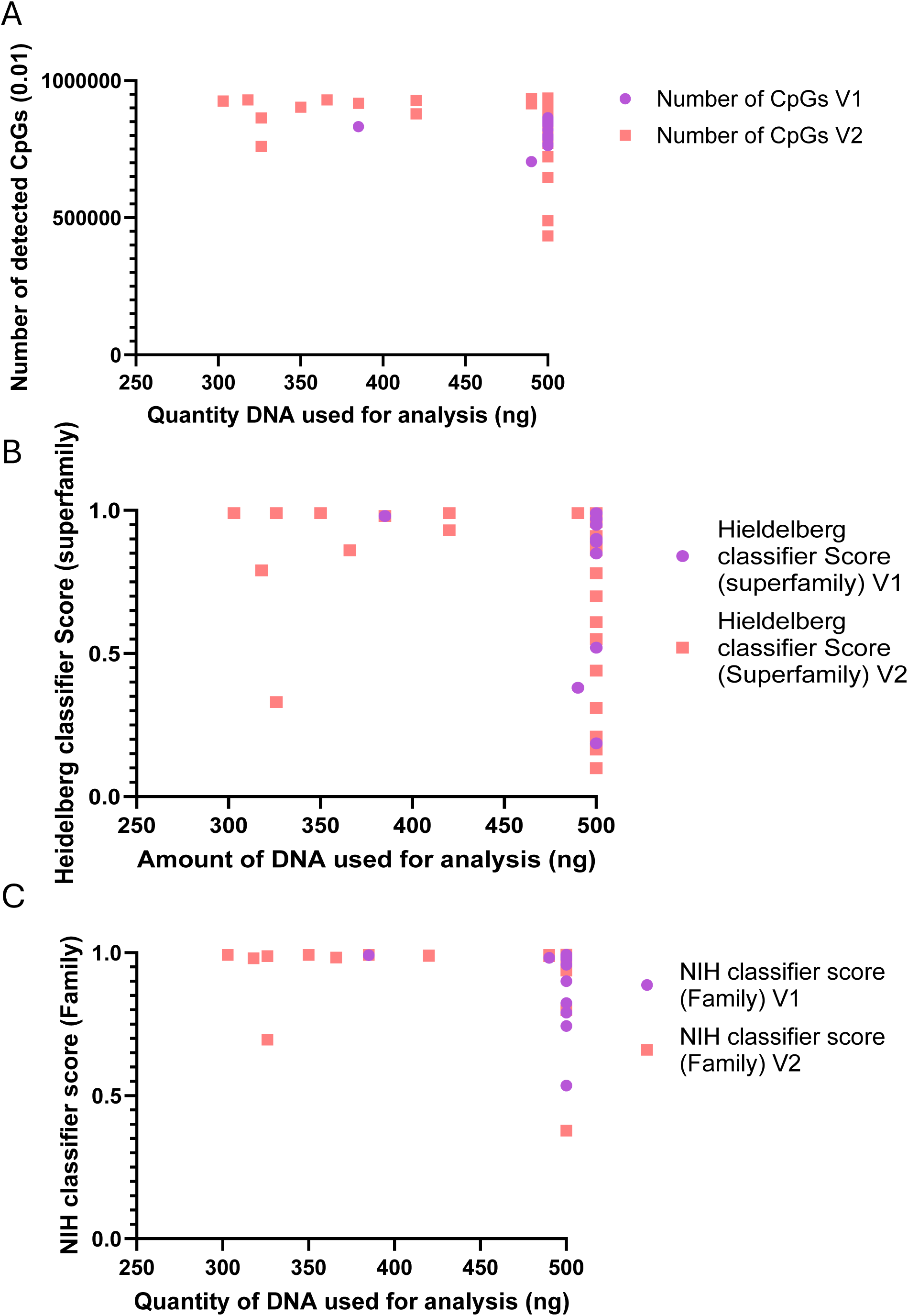
A. Dotplot of 88 samples (duplicates excluded) detectable CpGs against quantity of DNA used for analysis. Samples ran on V1 (33 samples) and represented in violet and samples ran on V2 represented in red. B. Dotplot of 88 samples (duplicates excluded) Heidelberg scores at the superfamily level against quantity of DNA used for analysis. Samples ran on V1 (33 samples) and represented in violet and samples ran on V2 represented in red. C. Dotplot of 88 samples (duplicates excluded) NIH methylscape scores at the family level against quantity of DNA used for analysis. Samples ran on V1 (33 samples) and represented in violet and samples ran on V2 represented in red.

**Supplementary figure 5.**
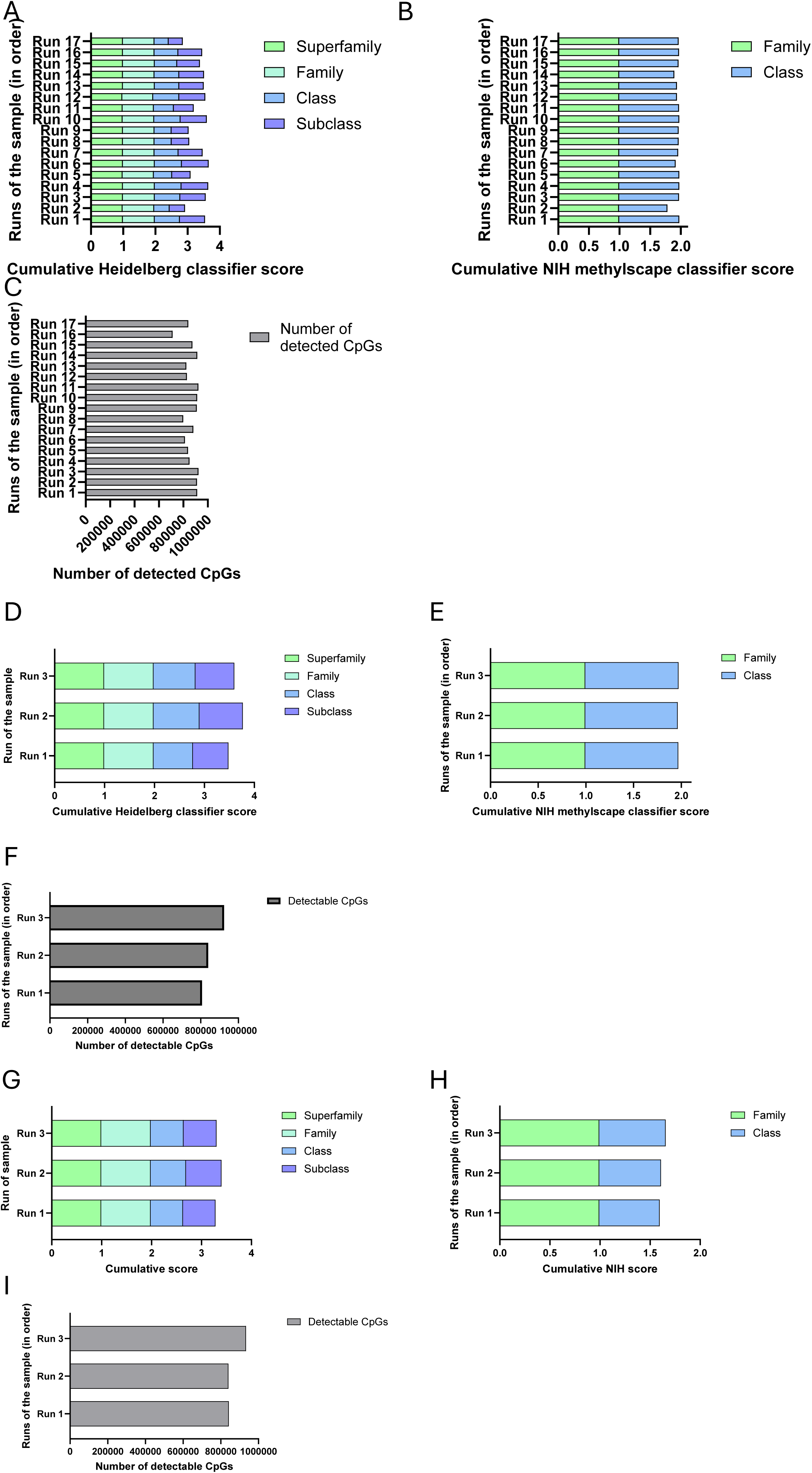
A, D, G. Cumulative Heidelberg classifier scores (superfamily (green), family (cyan), class (blue) and subclass (purple)) obtained for the first control sample ran 17 times (A), the second control sample ran 3 times (D) and the third control sample ran 3 times (G) (medulloblastoma) (in order of run). B, E, H. Cumulative NIH methylscape classifier scores (Family (green) and class (blue)) obtained for the first control sample ran 17 times (B), the second control sample ran 3 times (E) and the third control sample ran 3 times (H) (medulloblastoma) (in order of run). C, F, I. Number of CpGS detected for each of the runs of the same first control sample (medulloblastoma) (in order of run) (C), the second control sample (F) and the third control sample (I).

**Supplementary figure 6.**
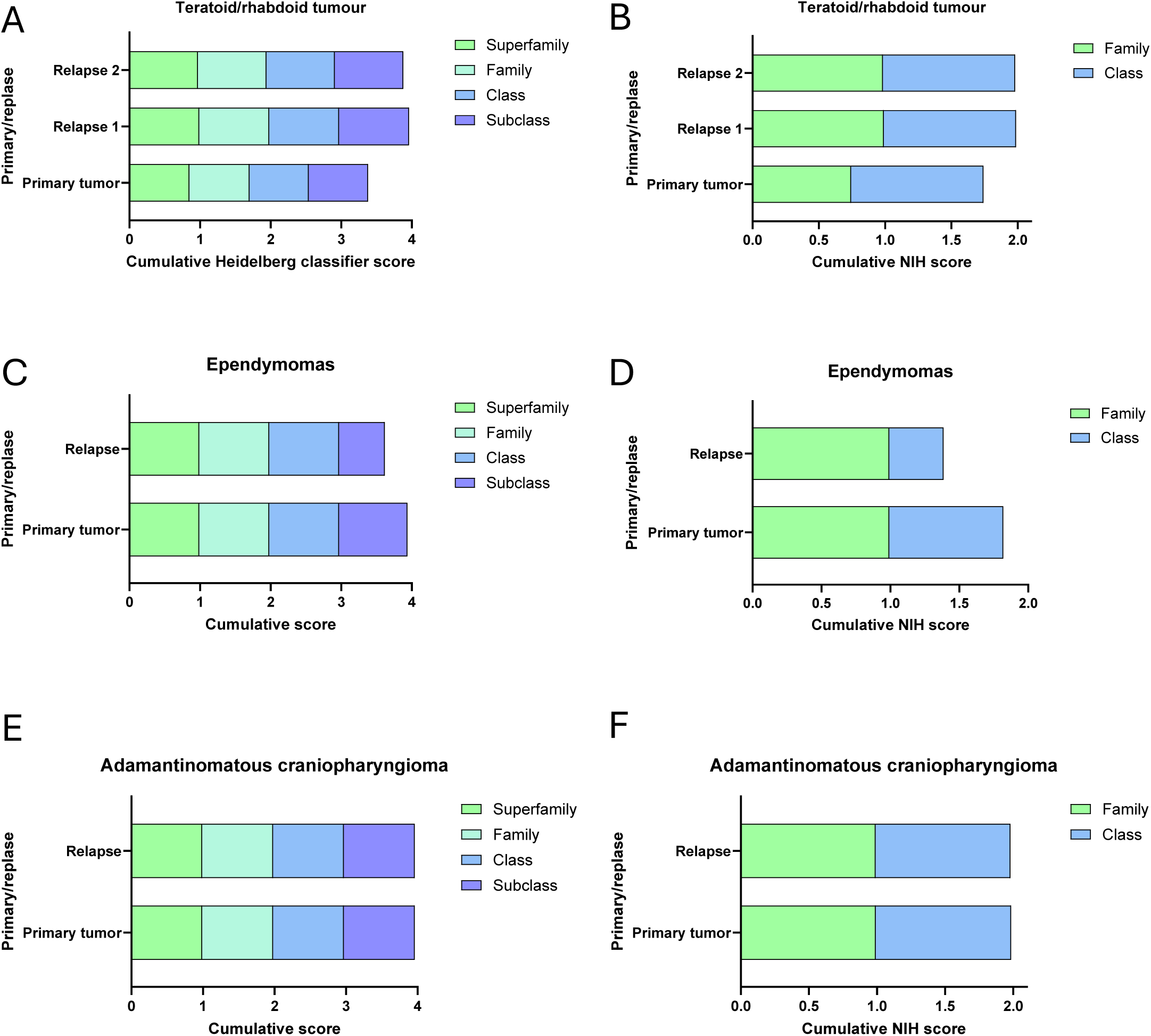
A, C, E. Cumulative Heidelberg classifier scores (superfamily (green), family (cyan), class (blue) and subclass (purpule)) obtained for the teratoid/rhabdoid tumor for primary tumor, first relapse and second relapse (A), the ependymomas samples ran for primary tumor, first relapse (B) and the adamantinomatous craniopharyngioma sample ran for primary tumor, first relapse. B, D, F. Cumulative NIH methylscape classifier scores (Family (green) and class (blue)) obtained for the teratoid/rhabdoid tumor for primary tumor, first relapse and second relapse (A), the ependymomas samples ran for primary tumor, first relapse (B) and the adamantinomatous craniopharyngioma sample ran for primary tumor, first relapse.

